# Longitudinal changes in DNA methylation associated with clozapine use in treatment-resistant schizophrenia from two international cohorts

**DOI:** 10.1101/2023.11.17.23298669

**Authors:** Amy L. Gillespie, Emma M. Walker, Eilis Hannon, Grant McQueen, Kyra-Verena Sendt, Alessia Avila, John Lally, Cynthia Okhuijsen-Pfeifer, Marte van der Horst, Alkomiet Hasan, Emma L. Dempster, Joe Burrage, Jan Bogers, Dan Cohen, Marco P. Boks, Alice Egerton, Jurjen J. Luykx, Jonathan Mill, James H. MacCabe

## Abstract

The second-generation antipsychotic clozapine is used as a medication for treatment-resistant schizophrenia. It has previously been associated with epigenetic changes in pre-clinical rodent models and cross-sectional studies of treatment-resistant schizophrenia. Cross-sectional studies are susceptible to confounding, however, and cannot disentangle the effects of diagnosis and medication. We therefore profiled DNA methylation in sequential blood samples (n=126) from two independent cohorts of patients (n=38) with treatment-resistant schizophrenia spectrum disorders who commenced clozapine after study enrolment and were followed up for up to six months. We identified significant non-linear changes in cell-type proportion estimates derived from DNA methylation data - specifically B-cells - associated with time on clozapine. Mixed effects regression models were used to identify changes in DNA methylation at specific sites associated with time on clozapine, identifying 37 differentially methylated positions (DMPs) (p < 5×10^−5^) in a linear model and 90 DMPs in a non-linear quadratic model. We compared these results to data from our previous epigenome-wide association study (EWAS) meta-analysis of psychosis, finding evidence that many previously identified DMPs associated with schizophrenia and treatment-resistant schizophrenia might reflect exposure to clozapine. In conclusion, our results indicate that clozapine exposure is associated with changes in DNA methylation and cellular composition. Our study shows that medication effects might confound many case-control studies of neuropsychiatric disorders performed in blood.

## Introduction

Schizophrenia is a complex and heterogeneous psychiatric disorder characterized by episodic psychosis, altered cognitive functioning, and often relatively persistent negative symptoms. Despite the significant contribution to the global burden of illness and decades of research, treatment options in clinical practice have not substantially changed over recent decades. Furthermore, the most commonly prescribed medications have limited effectiveness for approximately a quarter of patients^1^. These patients, commonly described as treatment-resistant^2^, are typically prescribed clozapine. Clozapine is a second-generation antipsychotic consistently shown by meta-analyses to have superior effectiveness over other antipsychotics, not only in treatment-resistant schizophrenia (TRS) but potentially in schizophrenia generally^3^. In meta-analyses of clinical trials^4^ as well as real-world settings^5^, clozapine has been associated with favourable outcomes compared to other antipsychotics in a range of patients and scenarios^6,7^. However, owing to its profile of adverse drug reactions – which includes frequent weight gain and sedation and increased risk of rare but potentially life-threatening possibilities such as agranulocytosis and myocarditis – clinical guidelines recommend that clozapine is reserved for TRS^8^. Unfortunately, over half of the patients starting a trial of clozapine still do not experience adequate symptom relief^1^ resulting in unnecessary exposure to a heavy adverse drug reaction burden. There is therefore a pressing need to understand the role of clozapine in schizophrenia in general and TRS in particular^9^.

While our understanding of the molecular changes in the brain that underpin the development of schizophrenia remains limited, we do know that the aetiology includes a strong genetic component^10–12^ in combination with numerous environmental risk factors^13^. There is increasing evidence that epigenetic processes - which regulate gene expression via modifications to DNA, histone proteins, and chromatin – might play a role in the aetiology of schizophrenia or may alter as a result of having schizophrenia^14–16^. DNA methylation (DNAm) is the best-characterized epigenetic modification, stably influencing gene expression via disruption of transcription factor binding and recruitment of methyl-binding proteins that initiate chromatin compaction and gene silencing. High-throughput profiling methods for quantifying DNAm across the genome have enabled researchers to perform epigenome-wide association studies (EWAS) aimed at identifying differences in DNAm at specific sites associated with disorders including schizophrenia^16^.

Our recent large-scale analysis of schizophrenia cases identified widespread differences in DNAm in blood samples from patients with schizophrenia compared to healthy controls^16^. Of note, we also found differences in a sub-group of patients who had been prescribed clozapine during their lifetime and found that many schizophrenia-associated DNAm differences were only present in patients with TRS; however, it is unclear whether these alterations were driven by characteristics of treatment-resistance or resulted from exposure to clozapine. Pre-clinical rodent studies have shown that clozapine alters histone acetylation and recruits DNA demethylation enzymes^17–19^. Furthermore, analyses of human patients have indicated that clozapine exposure is associated with significant changes in DNAm at specific sites across the genome, although these studies have either lacked in rigour (e.g. not controlling for common confounds such as cellular heterogeneity between individuals)^20^ or have been cross-sectional^16,21^.

The dynamic nature of epigenetic processes means that cross-sectional EWAS analyses can be confounded by many potential factors^22,23^. Additionally, cross-sectional studies obscure the temporal and causal relationship between DNAm changes and exposures of interest. This may be particularly pertinent in case-control analyses, where medication exposure is likely only applicable to cases and controls are medication naïve. This also applies to TRS, where clozapine exposure only occurs in this subset of cases and non-TRS schizophrenia patients will be prescribed other antipsychotics. This correlation between diagnosis and exposure is therefore impossible to resolve in a cross-sectional study. An alternative, powerful approach to attribute changes in DNAm to environmental exposure or diagnosis is to perform longitudinal profiling within the same individual.

In this study we quantified clozapine-associated changes in DNA methylation using a longitudinal EWAS design. We incorporated samples from two independent cohorts of patients with treatment-resistant schizophrenia spectrum disorders who started clozapine after study enrolment and baseline measurements. We identified changes in DNAm at specific sites following exposure to clozapine and used blood cell deconvolution approaches to identify changes in the proportion of different cell-types following exposure. We explored whether our clozapine-induced differences overlapped with those previously identified from cross-sectional case-control analyses of patients with schizophrenia, and those specifically with TRS. Our results show that clozapine exposure possibly explains some of the variation in DNAm previously attributed to a diagnosis of schizophrenia.

## Methods

### Participant recruitment and sample collection

We recruited participants from two European centres – Kings College London (KCL) in the United Kingdom and University Medical Center (UMC) Utrecht in the Netherlands. The inclusion criteria and sample processing protocols were equivalent between centres, as described below. Eligible participants were adult inpatients or outpatients (aged 18 or over), who were due to commence clozapine as part of their normal clinical care. Inclusion required that participants were clozapine naïve or had not taken clozapine for at least 3 months prior to the study. All participants were able to speak and read the local language. Participants with mental capacity to consent provided written informed consent to study procedures. At the KCL centre, the study was also open to participants lacking capacity to consent if a consultee advised assent on their behalf (see **Supplementary Methods**). All study procedures were conducted in accordance with the Declaration of Helsinki.

#### KCL (London) Cohort

Between August 2014 and December 2018, participants were recruited from inpatient and outpatient services within the South London and Maudsley/Oxleas NHS Foundation Trusts. Eligible participants met ICD-10 criteria for schizophrenia (F20) or schizoaffective disorder (F25). Inclusion criteria included at least two previous trials of a non-clozapine antipsychotic, each within the recommended dose range for at least six weeks, and a diagnosis of treatment-resistant schizophrenia provided by their psychiatrist. General exclusion criteria included drug dependency as primary disorder, as defined in DSM-IV, or pregnancy. Participants were offered financial reimbursement for their participation of £5 for each blood collection and £5 for each set of clinical assessments. This study was approved by London South East NHS ethics committee (Ref: 13/LO/1857).

#### UMC-U (Utrecht) Cohort

Between March 2016 and September 2019, participants were recruited from services in the Netherlands and Germany by the CLOZIN (CLOZapine INternational) consortium. Eligible participants were diagnosed with schizophrenia, schizophreniform disorder, schizoaffective disorder, or psychotic disorder not otherwise specified (according to DSM-IV or -V criteria) and a diagnosis of treatment-resistance. Exclusion criteria were involuntary admission to a psychiatric unit and a history of Parkinson’s disease. Participants were offered a financial reimbursement for participation in this study of €5 per study visit. Recruitment for all sites was approved by their respective local Institutional Review Boards.

#### Sample Collection

Across the six months of the study period, participants were invited to attend between three and five study visits (**Figure 1** **and Supplementary Methods**). Peripheral whole blood samples were collected at each study visit for DNAm analysis. All participants at both centres were required to attend a baseline visit before clozapine initiation, and were then invited to attend further visits if they remained on clozapine, including a final visit at 6 months. If they discontinued clozapine or withdrew consent to participate, they were withdrawn from the study. Only participants who provided blood samples for at least two visits (one before clozapine initiation and one after clozapine initiation) were included in the analysis.

**Figure 1.**
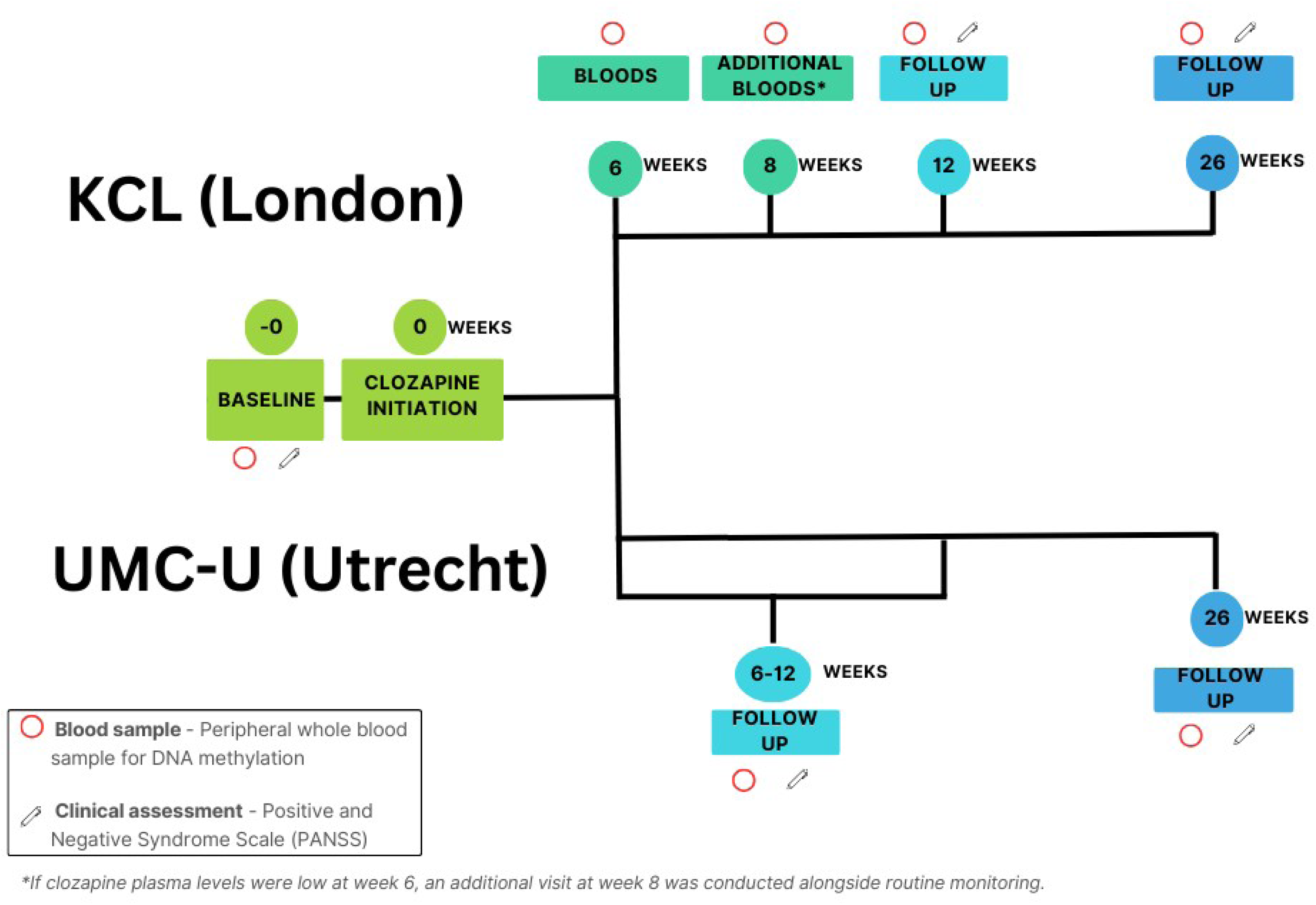
Time point chronology for each cohort. *For the KCL cohort, if clozapine plasma levels were low at week 6, an additional routine monitoring visit was conducted at week 8 and extra bloods were collected alongside this.

#### Clinical Assessment

For each participant we collected baseline demographic and clinical history through self-report and review of the patients’ medical records. The Positive and Negative Syndrome Scale (PANSS)^24^ was conducted at baseline, six-twelve weeks after baseline and six months after baseline, by trained researchers. To assess the efficacy of clozapine in both cohorts, we assessed the percentage change in PANSS total score from baseline to six-twelve weeks using the formula: (follow up score – baseline score) / baseline score × 100, after subtracting minimum possible scores at each time point. Participants were classed as responders to clozapine if they experienced a minimum of a 25% reduction in PANSS total between baseline and follow up, as recommended for TRS^25^.

### Sample processing

EDTA tubes with whole blood were delivered to the BRC Bioresource (KCL) or the Human Neurogenetics Unit (UMC-U) within three hours of collection. Samples were immediately stored on ice and then either processed for DNA extraction or stored at −20°C or −80°C before processing. For each sample, 1.5ug of DNA diluted to 25ng/ul was treated with sodium bisulfite treatment using the EZ-96 DNA methylation kit (Zymo Research, CA, USA). Genome-wide DNAm was profiled for the KCL samples using the Illumina Infinium HumanMethylation450 BeadChip (Illumina Inc, San Diego, California, United States) (“450K array”) and the UMC-U samples using the Illumina Infinium HumanMethylationEPIC BeadChip (Illumina Inc, San Diego, California, United States) (“EPIC array”). Arrays were organised so that all samples from the same individual were placed on the same array to minimise batch effects correlating with the outcome of interest. All samples were run on an Illumina iScan System (Illumina, CA, USA) using the manufacturers’ standard protocol in the Department of Clinical & Biomedical Sciences at the University of Exeter Medical School (https://www.exeter.ac.uk/departments/hls/clinical/).

### Data processing

All data processing and analyses were performed with R (v3.6.0). See **Figure 2** for an overview of the data processing steps used in this study.

**Figure 2.**
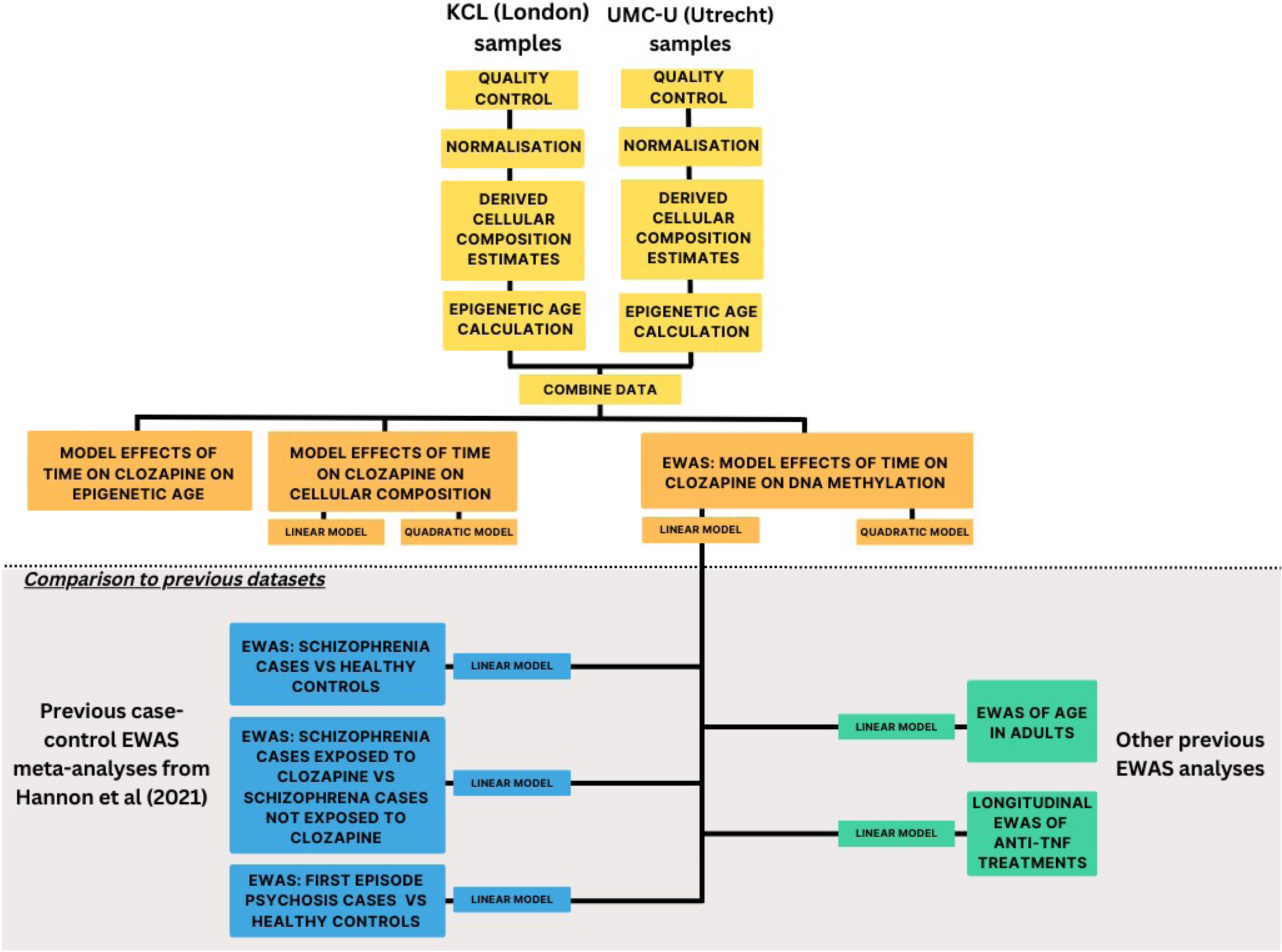
Overview of data processing and analysis steps.

#### Quality control (QC)

Data was imported from idat files and processed through a standard quality control pipeline^16^. This was done separately for each cohort as they were processed at different times using different versions of the Illumina BeadArray. This pipeline included the following steps: (1): checking methylated (M) and unmethylated (U) signal intensities and excluding samples with a median intensity < 2000 in either M or U, (2) calculating the bisulfite conversion statistic using the bscon function in the wateRmelon R package^26^, excluding any samples with <90, (3) principal component analysis of the DNAm data to confirm reported sex, (4) correlation of genotype data assayed by single nucleotide polymorphism (SNP) probes on each array (65 on the 450K and 59 on the EPIC array) to confirm samples from the same individual were genetically identical; and (5) use of the pfilter() function from the wateRmelon package to exclude samples with >1% of probes with detection P value > 0.05, probes with >1% of samples with detection P value > 0.05, and probes where greater than 5% of samples had a beadcount less than 3.

Following quality control, the KCL cohort had DNAm data at 448677 sites for 92 samples from 26 individuals and the UMC-U cohort had DNAm data at 802579 sites for 34 samples from 12 individuals. Normalization of the DNAm data was performed separately for each cohort using the *dasen* function in the *wateRmelon* package^26^.

#### Estimated proportions of blood cell types and epigenetic age

From these normalised DNAm data we derived several well-established epigenetic predictors, separately for each cohort. In light of known negative consequences of clozapine on blood cell counts^27–29^, we were interested in testing for changes in cellular composition during clozapine treatment. As cell count data were not available for a significant proportion of the DNA samples, we used a well-established reference-based algorithm to derive estimated proportions of blood cell types from the DNAm data using the Epigenetic Clock software^30^ which implements Houseman’s reference based constrained projection methodology^31,32^ for six cell types (CD8T, CD4T, natural killer cells, B cells, granulocytes and monocytes). Given most previous applications of epigenetic age have been in cross-sectional study designs, we were interested if there were progressive increases in epigenetic age that were proportional to the longitudinal time course of the study. Epigenetic age was therefore calculated using the clock developed by Zhang and colleagues^33^.

### Statistical analysis

Data for the two cohorts were merged into a single dataset for the 355,845 sites common to both, prior to statistical analysis.

To test for differences in cellular composition or epigenetic age during exposure to clozapine, a mixed effects regression model was fitted using functions from the lme4 and lmerTest R packages. Each of the six estimated cellular composition variables was modelled as the dependent variable (in a separate regression model) with time on clozapine (measured in days), age at baseline, and sex included as fixed effects and a random intercept term for participant to capture correlations between samples from the same individual and for institute (binary indicator). To allow for non-linear changes in DNAm as a function of time on clozapine we fitted a second set of quadratic models that additionally included time on clozapine squared (measured in days). Smoking has well known associations with DNAm at sites across the genome^34^. We therefore used a method developed by Elliot and colleagues^34^ to calculate a smoking score based on DNAm at these known sites. A mixed effects regression model was used to confirm there was no significant change in smoking score over time following clozapine exposure (P value = 0.58, **Supplementary Figure 1**).

#### Epigenome-wide association analysis (EWAS)

To test for differentially methylated sites associated with time on clozapine, the same mixed effects regression model, described above, was used, where DNAm level was modelled as the dependent variable. In this model, the six derived cellular composition variables were also included as fixed effect covariates. As with the cell composition analyses, we fitted both linear and quadratic models for time on clozapine.

A subsequent sensitivity analysis was conducted to test the effect of a sample collected at a notably later final time point. Running analyses both with and without this sample confirmed that the sample was not significantly influencing the results and therefore the results are presented with the sample included.

## Results

### Study overview and cohort characteristics

Schizophrenia spectrum patients about to commence treatment with clozapine were recruited from two European centres - King’s College London (KCL, n = 26 individuals) and University Medical Center Utrecht (UMC-U, n = 12 individuals). Participants had a mean age at baseline of 38.4 years (SD=12.5 years), were predominantly male (n =30; 79%), non-smoking (n = 27; 71%) with a primary diagnosis of schizophrenia (n = 28; 76%) and no prior use of clozapine (n = 32; 84%) (**Table 1**). Any prior clozapine use had been stopped at least three months before study initiation. The high rate of non-smoking patients was likely due to the restrictions on smoking for NHS in-patients in the KCL cohort. Patients had been prescribed a mean of 3.05 (SD=1.16) alternative antipsychotics prior to clozapine initiation. This is likely lower than average for a treatment-resistant sample, due to the study teams’ connections within specialist mental health services aimed at increasing prompt access to clozapine initiation (e.g. “clozapine clinics”). Both cohorts were comparable in terms of the proportion of male participants and mean number of previous antipsychotics, but the UMC-U cohort had no participants with prior exposure to clozapine and higher rates of baseline smoking.

**Table 1.** Summary of cohort demographics. Demographic statistics for KCL (Kings College London), UMC (University Medical Centre) and the two cohorts combined. N.B. PANSS scores not available for all participants. *These blood-sample only visits were only conducted for the KCL cohort, and the 8 weeks visit was only done for KCL participants if their clozapine plasma levels were low at 6 weeks (see Figure 1). ^1^One KCL participant had their baseline sample removed from analysis at the QC stage.

| Cohort |  | KCL | UMC | Combined |
| --- | --- | --- | --- | --- |
| Total Individuals |  | 26 | 12 | 38 |
| Total Samples |  | 92 | 34 | 126 |
| N samples at each time point | Baseline | 25<sup>1</sup> | 12 | 37 |
|  | 6 weeks* | 22 | n/a | 22 |
|  | 8 weeks* | 4 | n/a | 4 |
|  | 6-12 weeks | 21 | 12 | 33 |
|  | 26 weeks | 20 | 10 | 30 |
| Males | Individuals | 20 (76.9 %) | 10 (83.3%) | 30 (78.9%) |
|  | Samples | 70 (76.1 %) | 28 (82.4%) | 98 (77.8%) |
| Females | Individuals | 6 (23.1 %) | 2 (16.7%) | 8 (21.1%) |
|  | Samples | 22 (23.91 %) | 6 (17.6%) | 28 (22.2%) |
| Age at baseline (years) | Mean | 38.5 | 38.3 | 38.4 |
|  | SD | 11.7 | 14.5 | 12.5 |
| Number of antipsychotics prior to study | Mean | 3.3 | 2.6 | 3.1 |
|  | SD | 0.9 | 1.4 | 1.2 |
| Primary diagnosis of schizophrenia | Individuals | 20 (76.9%) | 8 (66.7%) | 28 (73.7%) |
|  | Samples | 74 (80.4%) | 23 (67.6%) | 97 (77.0%) |
| Clozapine naïve | Individuals | 19 (73.1%) | 12 (100%) | 31 (81.6%) |
|  | Samples | 72 (78.3%) | 34 (100%) | 106 (84.1%) |
| Non-smoker at baseline | Individuals | 20 (76.9%) | 7 (58.3%) | 27 (71.1%) |
| PANSS (total) at baseline | Mean | 79.0 | 80.5 | 79.5 |
|  | SD | 16.2 | 20.0 | 17.8 |
|  | Samples | 25 | 10 | 35 |
| PANSS (total) at 6-12 weeks | Mean | 62.8 | 64.7 | 63.4 |
|  | SD | 17.1 | 20.4 | 18.2 |
|  | Samples | 21 | 10 | 31 |
| Responders at 6-12 weeks | Individuals | 12 (57.1%) | 7 (70%) | 19 (61.3%) |
| PANSS (total) at 22-30 weeks | Mean | 61.9 | 62.7 | 62.2 |
|  | SD | 16.6 | 21.7 | 18.5 |
|  | Samples | 18 | 9 | 27 |

From these individuals, we quantified DNAm across the genome from a total of 126 blood samples using either the Illumina EPIC array (UMC-U samples) or Illumina 450K array (KCL samples) (see **Methods**). These longitudinal samples were collected over a follow up over a period of up to 6 months (**Figure 1**), with a mean of 3.32 timepoints per participant (SD=0.81) spanning a mean of 22.41 weeks (SD=7.51).

Mean PANSS score at baseline was 79.2 (SD=17.8), indicative of moderate illness (**Table 1**). Across both recruitment sites, the mean percentage reduction in PANSS was 31.67 (SD=34.33) (**Supplementary Figures 2A and 2B**) and 20 patients (60% of the 33 participants with available PANSS scores) were classified as treatment-responders at follow up, which is higher than typical within treatment-resistant schizophrenia^1^. Both cohorts were comparable in terms of mean baseline PANSS score and response rate to clozapine (**Table 1**).

### Accumulation of epigenetic age is slower than chronological age

We found a strong positive correlation between reported age and epigenetic age (*r* = 0.954, *p* = 8.07×10^−67^) across all samples, as expected. Whilst not significant (*p* = 0.300), we also found that the mean change in epigenetic age per study day was positive and equivalent to an increase of 0.622 years per chronological year (**Supplementary Figure 3**). This is consistent with previous reports showing that the epigenetic age of older individuals is underestimated, potentially reflecting the miscalibration of aging effects with a slower increase in epigenetic age relative to chronological age.

### Estimated proportion of B-cells changes during early clozapine exposure

Using a mixed-effects regression model, we found that time on clozapine was not linearly associated with changes in the estimated proportion of any of the cell types tested (**Supplementary Table 1)**. We additionally tested for a non-linear relationship between time on clozapine and cellular proportions by adding a quadratic term to the regression model to capture more acute changes that stabilise or even reverse during the study period (**Supplementary Table 1**). From a comparison of regression model fit, we observed that - for the estimated proportions of B cells - the quadratic model was a better fit to the data (linear AIC −583.26, quadratic AIC −598.44, *p* = 3.39×10^−5^). Visualising the relationship between B cell proportion and time on clozapine, there was significant individual variability, but the mean proportion of B-cells showed an initial acute decrease until 14 weeks before increasing (**Figure 3**). By 28 weeks, the mean proportion of B-cells was equivalent to the proportion at baseline. No other significant changes were detected for any other estimated cell type.

**Figure 3.**
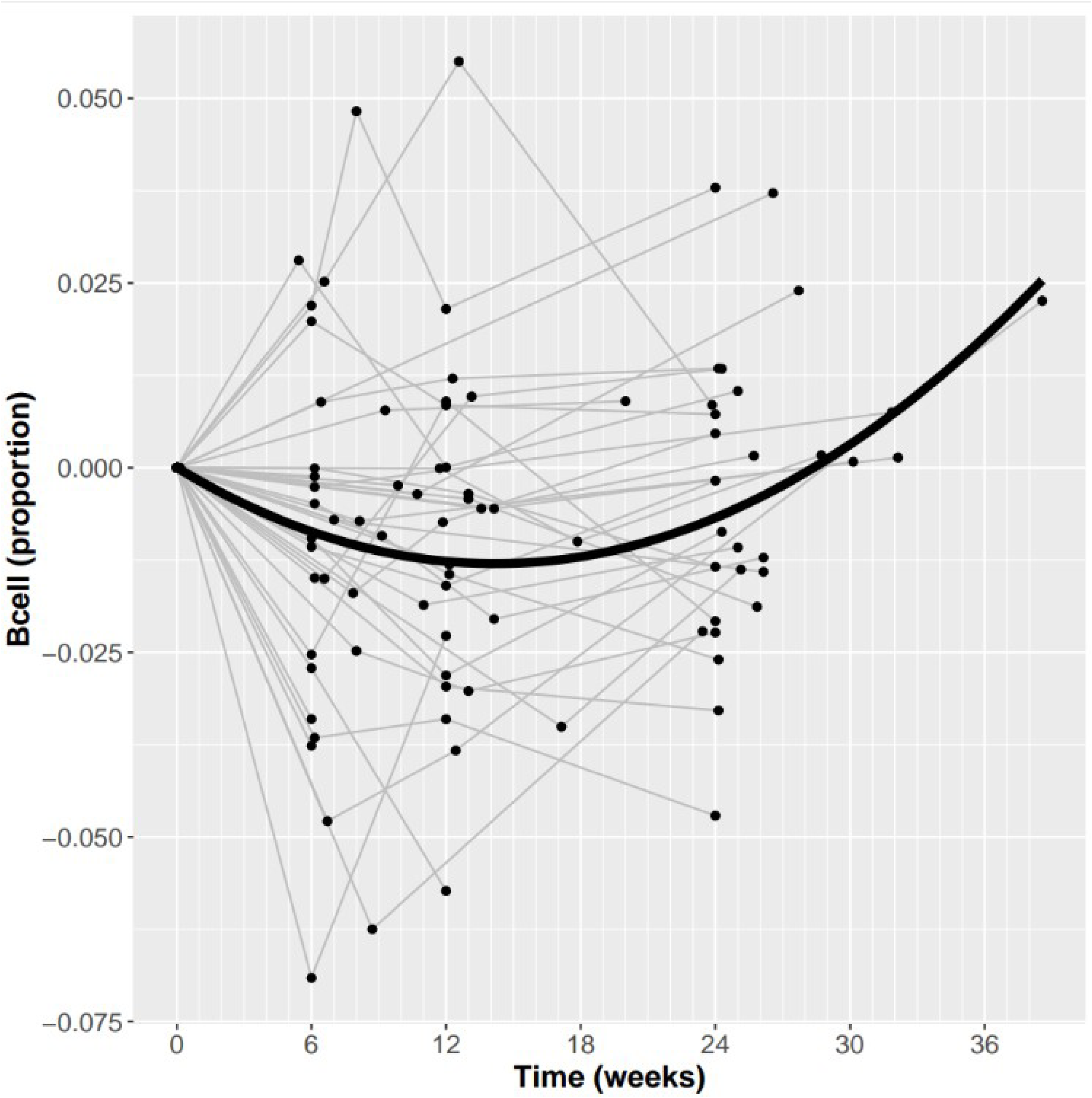
Non-linear change in proportion of B cells during exposure to clozapine. Plotted is the estimated proportion of B cells derived from DNA methylation data using the reference-based Houseman deconvolution algorithm (y-axis) against time on clozapine (x-axis; weeks). Each line represents a patient, where data points have been standardized by the patients baseline value, such that all lines start at the origin of the graph. The bold line represents the estimated model fit from the regression analysis. Note one patient was excluded from this plot has they had no baseline sample.

### Differential DNAm associated with time on clozapine

#### Linear regression model

To identify differentially methylated positions (DMPs) in blood associated with clozapine treatment, we performed an epigenome-wide association study (EWAS) using the same mixed effects regression model described above, with the addition of the derived cellular composition variables as covariates. Firstly, testing for linear effects of clozapine exposure against DNAm, we identified one DMP at an experiment-wide significance threshold (*p* < 1×10^−7^) with 37 DMPs at a more relaxed “discovery” threshold (*p* < 5×10^−5^; **Supplementary File 1**). The top significant DNAm site was cg06831576 (mean change in DNAm per day on clozapine = −1.25×10^−4^, *p* = 6.24×10^−8^, **Figure 4A**) located upstream of CDH8 on chromosome 16, which was characterized by a significant decrease in DNAm over the study period. Using public data in the EWAS Catalog we found that DNAm at this site has previously been associated with smoking^35^ and age in a childhood cohort^36^ (with an opposite direction of effect) and was also associated with circulating IL2RB protein levels^37^.

**Figure 4.**
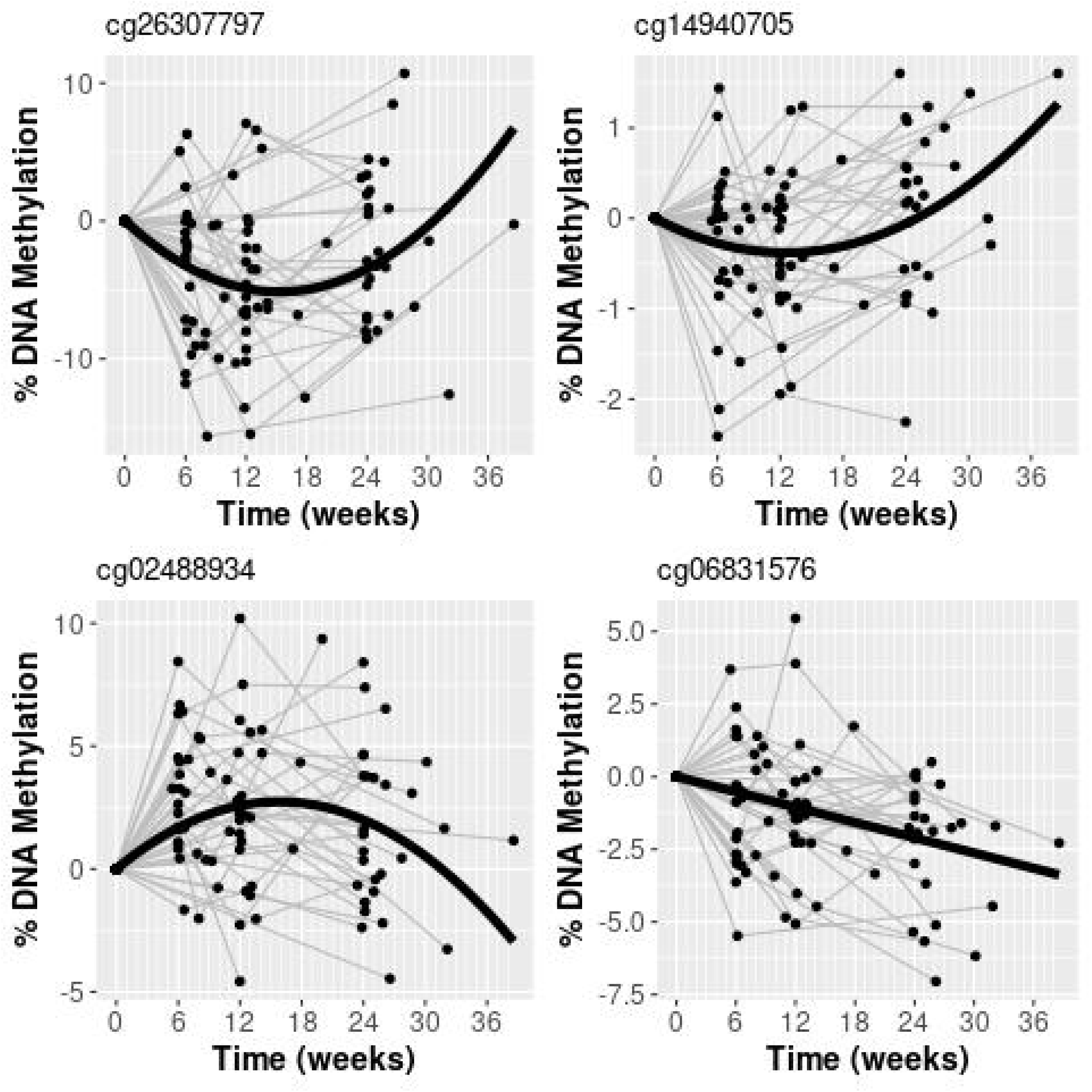
Change in methylation during clozapine exposure for notable probes. Each line represents a patient, where data points have been standardized by the patients baseline value, such that all lines start at the origin of the graph. **a)** cg0683157 - The top significant DNA methylation site from the linear mixed effects regression model. **b)** The top significant DNA methylation site from the non-linear mixed effects regression model. **c** & **d)** significant methylation sites from the non-linear mixed effects model previously associated with schizophrenia

#### Comparison to other longitudinal datasets

While our within-individual longitudinal study design enabled us to control for inter-individual confounders associated with clozapine exposure, all our participants were also all aging at the same chronological rate in parallel to all being treated with clozapine. Therefore, it is possible that our study design could detect age-associated DMPs, and it would be challenging to determine whether age or clozapine exposure is the cause of any identified DMPs. To explore this further, we looked up previous reported associations for the additional 36 discovery DMPs. While 14 and 1 DMPs were also associated with age in childhood, and prenatal samples respectively^36,38^, none of our discovery DMPs were associated with age in adults. Of note, if we look at age-associated DMPs from a large EWAS of age performed in adults (n = 656)^39^, there is a depletion of consistent directions of effect with our DMPs (1570 of 3726, 42%, *p* = 7.74×10^−22^) (**Supplementary Figure 4**). In addition, we also compared our results to another longitudinal study looking at effects of anti-TNF treatments on DNAm in IBD patients^40^. Both studies are potentially confounded by participants’ aging, but the medication effects should be very different. Indeed, when taking our 37 DMPs we found no evidence of consistent effects across our study and the IBD study (699 of 1591, 43.93%, sign test *p*=0.99) (**Supplementary Figure 5**), supporting the interpretation that our results are not confounded by the aging process and are attributable to clozapine.

#### Quadratic regression model

Again, we tested for non-linear effects of clozapine exposure against DNAm using a quadratic model. We identified one significant DMP at an experiment-wide significance threshold, where the quadratic model was found to be a better fit to the data than the linear model. An intergenic DNAm site on chromosome 11 (cg26307797) was associated with an average initial acute decrease in DNA methylation until 15 weeks, before which DNA methylation increased again (mean change in DNAm per day on clozapine = −9.44×10^−4^, *p* = 1.81×10^−8^, mean change in DNAm per day^2^ on clozapine = 4.39×10^−6^, *p* = 6.77×10^−8^) (**Figure 4B**). A further 89 DNAm sites were non-linearly associated with clozapine exposure at our discovery threshold, exhibiting acute changes that then normalised, in both directions (**Supplementary File 2**). 49 DMPs initially decreased following clozapine initiation, before increasing again while 40 DMPs increased before decreasing. In the majority of cases, the inflection point occurred at 14-25 weeks (**Supplementary Figure 6**, mean = 14.7 weeks, SD=1.80), meaning that the average DNAm level had returned to baseline levels by the end of the study period for these sites. Of note, two DMPs (cg14940705, annotated to SGK3, and cg02488934, intergenic on chromosome 1) (**Figures 4C,4D**) have been previously associated with schizophrenia.

### DNAm changes induced by clozapine exposure are consistent with those associated with schizophrenia in a large blood-based EWAS

Given the overlap between our clozapine-associated DMPs and previous schizophrenia-associated DMPs, we were interested in harnessing these data to aid the interpretation of previous cross-sectional EWAS analyses of schizophrenia which might be confounded by the medication status of patients. Leveraging the results of a previous large meta-analysis of schizophrenia cases^16^, we explored whether schizophrenia-associated variation might reflect clozapine use.

#### Comparison to schizophrenia case-control

From our previous EWAS of schizophrenia cases (n = 2,379) and controls (n = 2,104)^16^, we took the 848 significant DMPs that overlapped with sites assessed in our combined clozapine dataset and compared effect sizes, using the estimated changes in DNAm from the linear model of time on clozapine. We identified highly consistent effects, with 715 (84.3%) of schizophrenia DMPs being characterised by the same direction of effect in our study of clozapine exposure, a significantly higher proportion than expected by chance (*p* = 2.34×10^−97^, **Figure 5A**). This is consistent with a previous analysis of these schizophrenia-associated DMPs, reporting enrichment with associations from a cross-sectional study looking at differences within schizophrenia cases comparing those prescribed clozapine and those prescribed other anti-psychotics. Of note the majority of these (652/848, 76.9%) sites were hypermethylated in patients prescribed clozapine.

**Figure 5.**
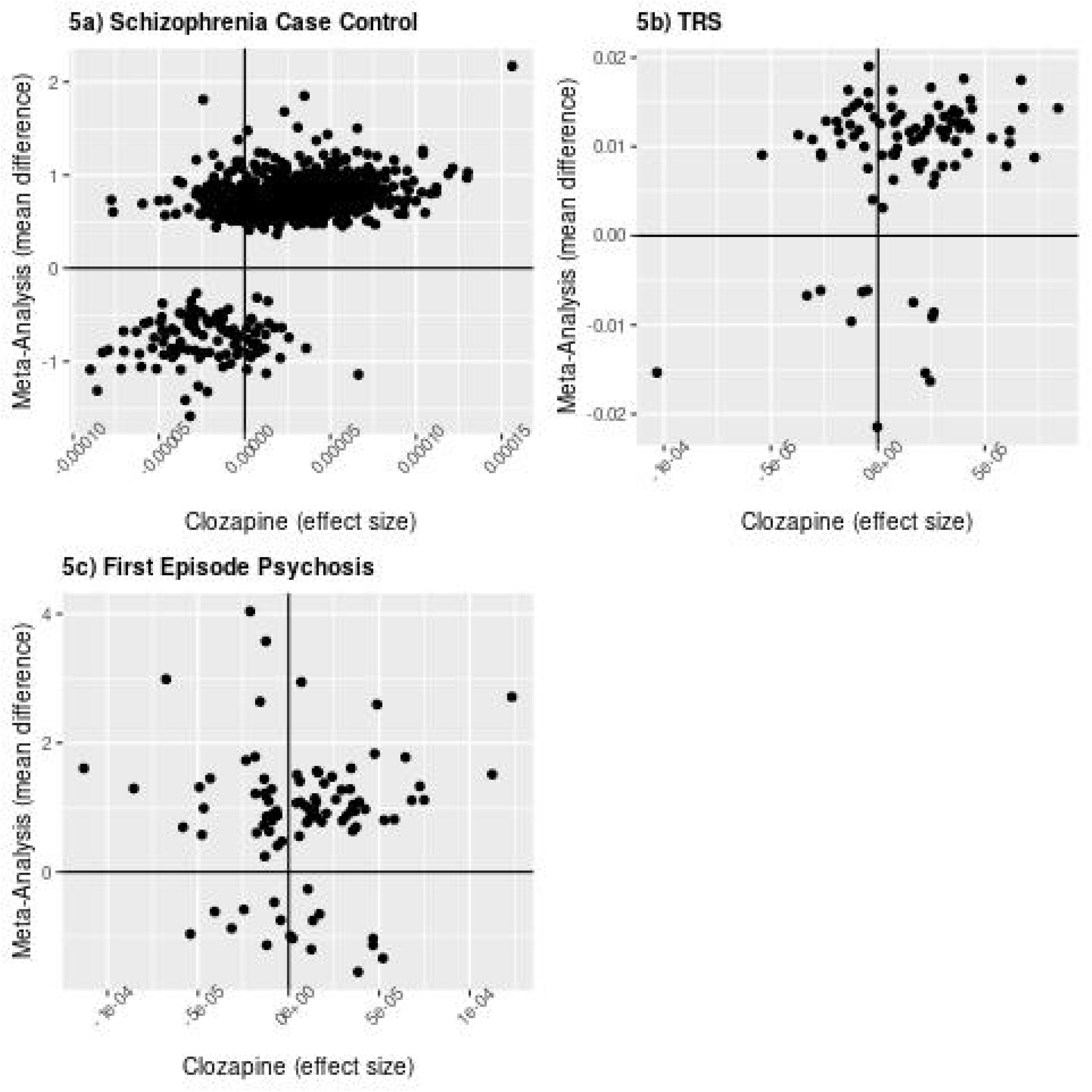
Overlap between probes assayed in Hannon et al (2021) meta-analysis and our combined analysis. **a)** The 848 identified DMPs in the Hannon et al. (2021) analysis of schizophrenia cases and controls. **b)** The top 100 probes in the Hannon et al., (2021) analysis of treatment-resistant schizophrenia cases and non treatment-resistant schizophrenia cases. **c)** The top 100 probes in the Hannon et al., (2021) analysis of first episode psychosis cases and controls.

#### Comparison to clozapine-exposed schizophrenia case-control

Next we compared our results from longitudinal exposure to clozapine with the cross-sectional within-schizophrenia EWAS of clozapine treatment^16^, which compared DNAm between 399 schizophrenia cases with lifetime exposure to clozapine and 636 schizophrenia cases never exposed to clozapine, focusing on the top 100 most statistically significant sites. Of these, 90 overlapped with sites included in our final EWAS dataset. Of these, we found 62 (68.9%) were associated with the same direction of effect, again a significantly higher proportion than expected by chance (*p* = 2.19×10^−4^) (**Figure 5B**). This suggests that many of the differences identified in the cross-sectional analysis potentially reflect clozapine exposure rather than factors related to being treatment-resistant per se.

#### Comparison to first-episode psychosis case-control

Finally, to check the specificity of these results, we also compared our results to DNAm differences associated with first episode psychosis, from an analysis comparing DNAm between 698 first episode psychosis cases and 724 controls^16^. Of the 100 top-ranked DMPs, 91 overlapped with sites assessed in the current study with no significant consistency in the direction of effect (*p* = 0.15) (**Figure 5C**). This strengthens the interpretation that the DNAm changes identified in our analyses are likely to be attributable to clozapine exposure, rather than consequences of non-specific psychosis risk factors.

## Discussion

We report the most extensive study of DNA methylation changes associated with clozapine use, profiling sequential blood samples from two cohorts of patients with treatment-resistant schizophrenia over the course of 6 months exposure to clozapine. We identify within-participant changes in derived cell proportion estimates and find differentially methylated positions (DMPs) associated with duration of clozapine use. By comparing our results with those from a previous large EWAS meta-analysis of schizophrenia, we demonstrate the value these longitudinal clozapine-exposure data have for interpreting previous case-control analyses of psychiatric disorders. Our analyses clarify and build upon the findings reported by Hannon and colleagues^16^ that many differences identified in blood EWAS analyses of schizophrenia are likely driven by data from patients prescribed clozapine, and therefore likely attributable to medication effects rather than the diagnosis of treatment-resistant schizophrenia *per se*.

A prominent clinical concern regarding clozapine treatment is the risk of agranulocytosis, whereby white blood cells are dramatically reduced ^28,29,41^. In light of this, the changes we report in estimated cellular composition are interesting. It might have been expected that we would see changes in the proportion of granulocytes, however clozapine-induced life-threatening agranulocytosis is very rare^42,43^, and mandatory weekly blood monitoring means that any clinically significant drop in neutrophils would have been identified, and led to clozapine discontinuation and withdrawal from the study. Instead of changes in granulocytes, we report a small but significant acute change in the proportion of B cells in our cohorts, with an average initial decrease and subsequent increase at 14 weeks back to baseline levels by 28 weeks. Interestingly, a separate analysis of the participants in the KCL cohort reported that clozapine exposure was associated with a reduction in participants’ immunoglobulin levels at 12 and 24 weeks (and that greater reductions were associated with improved response to clozapine)^44^. Hannon and colleagues^16^ also reported changes in cellular composition in patients prescribed clozapine, although these associations were with proportions of other cell types, namely increases in the proportion of monocytes and granulocytes and decreases in the proportion of CD4+ T cells and CD8+ T cells. These differences may be explained by the limitations of derived cellular proportions or differences in study population and duration of clozapine exposure.

Some of the specific sites where we identified potential changes in DNAm associated with time on clozapine have previously been associated with schizophrenia. From our linear analysis, two DMPs have been previously associated with schizophrenia (cg01300684, annotated to SCOC and cg11820931, annotated to DDX21)^16^. In both cases, the sites were associated with hypermethylation in schizophrenia and hypermethylation after exposure to clozapine in our EWAS. From our non-linear analysis, another two DMPs have been previously associated with schizophrenia, cg02488934 which is intergenic on chromosome 1, and cg14940705, which is annotated to SGK3^16,45^. While both DMPs were hypermethylated in schizophrenia, they displayed opposing average trajectories with clozapine in our sample. This suggests that while medication may be the driving factor behind the previous schizophrenia association at these sites, drug effects may not always manifest in a predictable, easy to interpret manner.

Our EWAS analyses implicate a number of interesting genes that may be relevant to the mechanisms of clozapine. For example, results from our linear model implicated CDH8, a gene which encodes cadherin 8 (a protein which mediates calcium-dependent cell-cell adhesion, is expressed in brain and has been associated with autism and cortico-striatal circuits^46^). Results from our quadratic model implicated SGK3, a gene which encodes a serum- and glucocorticoid-inducible kinase (expressed in brain and previously associated with memory consolidation^47^), S100B, a gene which encodes a calcium-binding protein (highly expressed in glial cells and previously associated with dysfunctional calcium signalling in schizophrenia^48^), and C9orf4, a gene, which encodes a component of an AMPA receptor protein in the brain (a pathway involving glutamatergic signalling and found to be dysfunctional in schizophrenia^49^). However, the majority of these were only significant at a discovery threshold, so must be confirmed by further research.

Cross-sectional epigenetic studies are not optimal for identifying causal risk factors and are potentially influenced by environmental confounders. To circumvent these caveats our longitudinal design enabled us to profile within-participant changes in DNAm following clozapine initiation and correlated with exposure over time. Unfortunately, recruiting and retaining clinical participants for these studies is challenging; it involves their participation during a period of their illness when they are often the most acutely unwell, right before commencing a new antipsychotic. This meant we only had access to a relatively small number of patients, limiting our power to detect novel associations. However, we leveraged the power of our previous large case-control study of schizophrenia to show that clozapine exposure potentially acts as a confounder in the identification of disease-associated DMPs. DNAm differences identified in treatment-resistant schizophrenia patients are likely mediated by clozapine exposure rather than factors related to being treatment resistant per se. Our identification of non-linear changes in DNAm further suggests that such drug effects might be varied and complex, in way that are difficult to capture within case-control studies.

Our analysis assumes that the critical factor changing over the course of the study was exposure to clozapine, although our participants were also potentially exposed to multiple other environmental factors that may affect DNAm in a cumulative manner (e.g. diet, alcohol, smoking, or changes secondary to symptom improvement) and previously prescribed antipsychotic will have been cross-titrated with clozapine in many cases. With the absence of a control arm, we compared our data to another longitudinal study of a different type of medication and found little overlap, reassuring us that our results did not simply reflect changes associated with aging. Nonetheless, future studies should include a control arm to strengthen the conclusions that can be made from longitudinal data. Over half of our sample showed a clinical response to clozapine, higher than with previous literature^1^, but there was significant heterogeneity in symptom change over the study and – as mentioned above – our study lacked the statistical power to explore differences in DNAm associated with clinical response. Finally, it is well established that individuals metabolise clozapine differently. Our analyses may have been better powered if we were able to analyse clozapine level directly, but unfortunately, these data were not routinely available in the clinical records for these patients.

In conclusion, our analyses indicate that clozapine exposure is associated with changes in DNA methylation and estimated cellular composition over the first six months of exposure, and that these changes likely drive many of the identified changes in DNA methylation previously reported in case-control studies of schizophrenia and treatment-resistant schizophrenia.

## Data Availability

All data produced are available online. GEO acession number GSE237561

## Acknowledgements

This work was funded by the Medical Research Council, UK, Grant MR/L003988/1 to **A.E**. and by the European Community’s Seventh Framework Programme (FP7/2007–2013), grant 279227 to **J.H.M**. This work was also supported by grants from the UK Medical Research Council (MRC; MR/K013807/1 and MR/R005176/1) to **J.M**. High-performance computing was supported by MRC Clinical Research Infrastructure Funding (MR/M008924/1) to **J.M**. This study presents independent research funded in part by the NIHR Maudsley Biomedical Research Centre at South London and Maudsley NHS Foundation Trust and King’s College London. This work was also supported by the National Institute for Health and Care Research Exeter Biomedical Research Centre and Oxford Health Biomedical Research Centre. The views expressed are those of the author(s) and not necessarily those of the NIHR or the Department of Health and Social Care. This study was endorsed by DZPG (German Center for Mental Health) (to **A.H**., FKZ: 01EE2303C). This work was supported by a personal Rudolf Magnus fellowship award to **J.J.L** (H-150) from the University Medical Centre Utrecht, Utrecht, The Netherlands. **A.A.** was supported by Fundação para a Ciência e a Tecnologia, scholarship ID SFRH/BD/115916/2016.

## Disclosures

Unrelated to this work, A.L.G has received consultancy fees from Zogenix. A.H was member of advisory boards of Boehringer-Ingelheim, Lundbeck, Janssen, Otsuka, Rovi and Recordati and received paid speakership by these companies as well as by AbbVie and Advanz. He is editor of the German schizophrenia guideline. All other authors have nothing to disclose.

## Notes

### Author Declarations

London South East NHS ethics committee gave ethical approval for this work (Ref: 13/LO/1857)

