## SupplementaryMaterials for "Longitudinal changes in DNA methylation associated with clozapine use in treatment-resistant schizophrenia from two international cohorts"

**Supplementary Materials**

**Supplementary File description**

**Supplementary File 1.** Mixed effects regression analysis results for the 37 discovery DMPs exhibiting a significant (P < 5x10-5) linear change in DNA methylation during exposure to clozapine. (qMoD = quadratic model)

**Supplementary File 2.** Mixed effects regression analysis results for the 89 discovery DMPs exhibiting a significant (P < 5x10-5) non-linear change in DNA methylation during exposure to clozapine. (qMod) = quadratic model

**Supplementary Methods**

*Consultee Procedures (KCL)*

The consultee was defined as a person non-professionally involved in caring for the patient or concerned with their welfare (normally their next-of-kin family member). They were advised on the role of the consultee, provided with a consultee information sheet and invited to ask any questions about the study before advising on the patient’s behalf. If there was any indication from the patient, consultee, clinical team, or anyone else involved in the patient’s care that the patient would not wish to participate, they were not enrolled in the study. During study participation, study researchers maintained contact with the consultee and clinical team. If the patient expressed any objections to the study or wishes to withdraw, they were withdrawn from the study immediately.

*Study Visit Schedule*

KCL participants were invited to attend a visit six weeks after clozapine initiation, a visit twelve weeks after clozapine initiation, and the final visit six months after clozapine initiation. Clozapine plasma levels were assessed at six week visit, to confirm a therapeutic dose of at least 350ng/mL. If clozapine plasma levels were below 350ng/mL, clinical review was conducted and participants had their clozapine plasma levels assessed approximately a fortnight later to confirm therapeutic dose had been reached. In the KCL participants this applied to, where possible, an additional blood sample for DNAm analysis was taken at this assessment (i.e. eight weeks after clozapine initiation).

UMCU participants were invited to attend a visit between 6-12 weeks after initiation, and the final visit 6 months after clozapine initiation. Clozapine plasma levels were assessed during clinical care and the second visit (between 6-12 weeks) was scheduled after the treating clinician determined they were at a stable dose.

When participants were unwilling or unable to attend a visit according to the study schedule, they were invited for a visit as close to this date as possible. If participants were unable or unwilling to attend one of the visits between baseline and final visit, they were still invited for subsequent visits.

**Supplementary figures and tables (7 in total)**

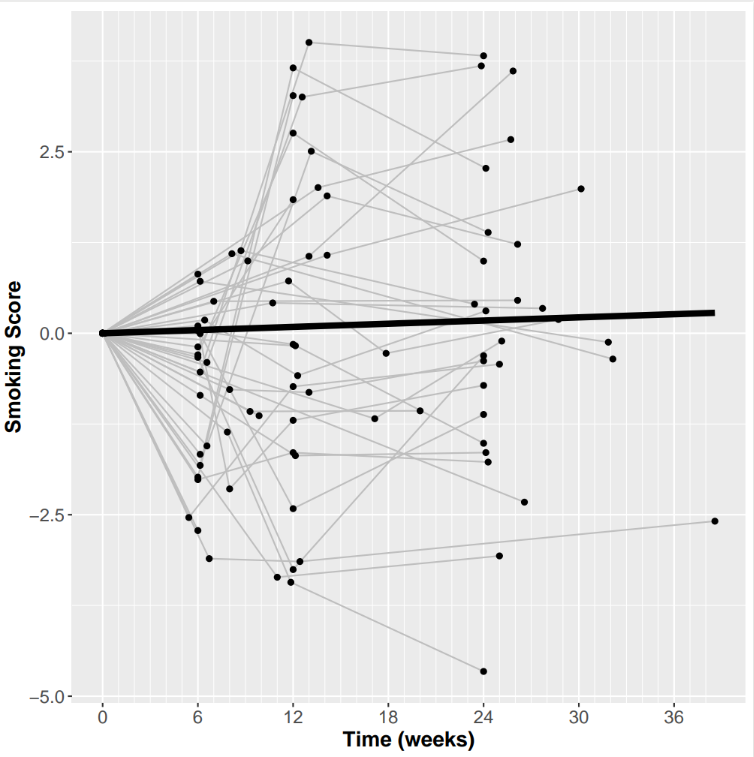

**Supplementary figure 1. Change in smoking score during clozapine exposure**. Plotted is the estimated smoking score derived from DNA methylation data using the method developed by Elliot et al. (2014) (y-axis) against time on clozapine (x-axis, weeks). Each line represents a patient, where data points have been standardized by the patients baseline value, such that all lines start at the origin of the graph. The bold line represents the estimated model fit from the regression analysis. Note one patient was excluded from this plot as their baseline sample was removed during QC.

**Supplementary Figure 2a**

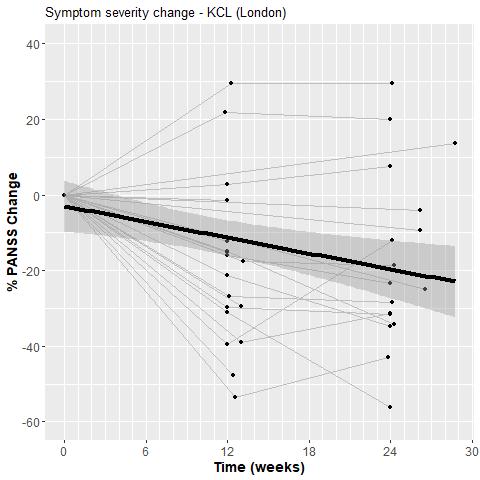

**Supplementary Figure 2b**

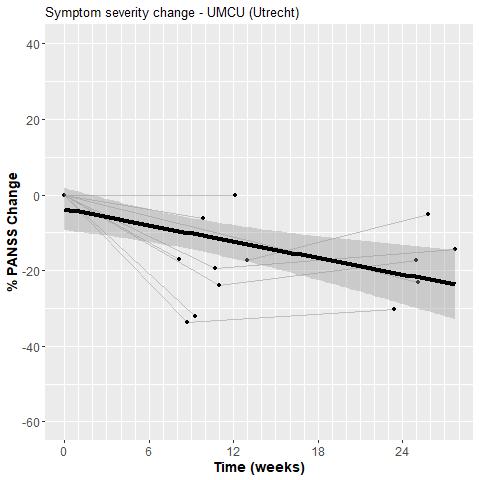

**Supplementary figure 2. Change in symptom severity during clozapine exposure**. **a) Symptom severity change in the KCL cohort. b) Symptom severity change in the UMCU cohort.** Plotted is the percentage change in PANSS score (y-axis) against time on clozapine (x-axis, weeks). Each line represents a patient, where data points have been standardized by the patients baseline value, such that all lines start at the origin of the graph. The bold line represents the estimated model fit from the regression analysis. NB: Only participants with PANSS scores available at baseline and follow-up were plotted

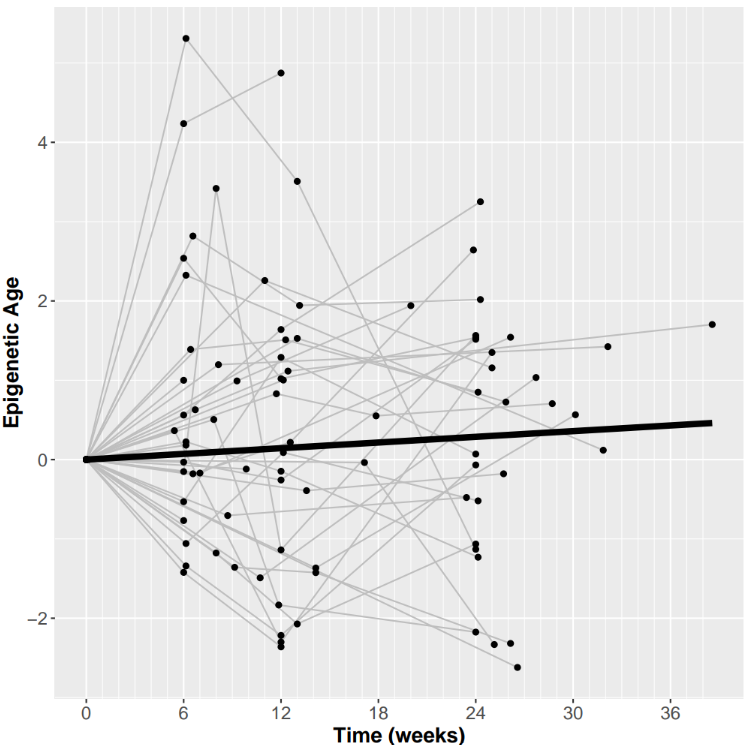

**Supplementary figure 3. Change in epigenetic age during clozapine exposure**. Plotted is the estimated epigenetic age derived from DNA methylation data using the methodology developed by Zhang (2019) (y-axis) against time on clozapine (x-axis, weeks). Each line represents a patient, where data points have been standardized by the patients baseline value, such that all lines start at the origin of the graph. The bold line represents the estimated model fit from the regression analysis. Note one patient was excluded from this plot has they had no baseline sample.

| **Cell Type** | **time^2^ Effect Eize** | **time^2^ SE** | **time^2^ P value** | **time Effect Size** | **time SE** | **time P value** | **ANOVA_Pr(>Chisq)** |
| --- | --- | --- | --- | --- | --- | --- | --- |
| CD8T | 9.03E-07 | 3.56E-07 | 1.30E-02 | -2.96E-05 | 2.38E-05 | 2.17E-01 | 1.30E-02 |
| CD4T | 1.59E-06 | 6.28E-07 | 1.32E-02 | -3.79E-05 | 4.19E-05 | 3.69E-01 | 1.32E-02 |
| NK | -2.68E-07 | 4.74E-07 | 5.73E-01 | -7.30E-07 | 3.06E-05 | 9.81E-01 | 5.72E-01 |
| Bcell | 1.33E-06 | 3.05E-07 | 3.44E-05* | -1.05E-05 | 2.16E-05 | 6.28E-01 | 3.39E-05 |
| Gran | -3.10E-06 | 1.21E-06 | 1.18E-02 | 7.10E-05 | 8.05E-05 | 3.80E-01 | 1.15E-02 |
| Mono | -6.01E-07 | 3.86E-07 | 1.23E-01 | 1.20E-05 | 2.53E-05 | 6.36E-01 | 1.24E-01 |

**Supplementary Table 1. Mixed effects regression analysis results for change in proportion of blood cell types during exposure to clozapine.** The estimated proportion of 6 different blood cell types (CD8T, CD4T, Natural Killer Cells, B cells, Granulocytes and Monocytes) were derived from DNA methylation data using the reference-based Houseman deconvolution algorithm. Results from the linear and quadratic model are shown.

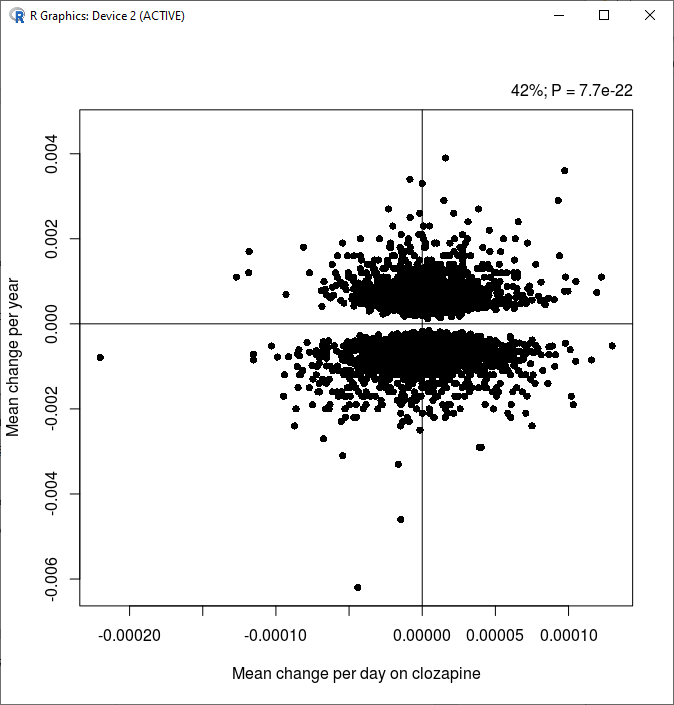

**Supplementary figure 4** – **Scatterplot of age associated differentially methylated positions (n = 3726) comparing the estimated effect of clozapine exposure (x -axis) and age (y-axis).** Age associated DMPs were identified from Battrum et al. (P < 1x10^-7^) and data extracted for these from the EWAS catalogue.

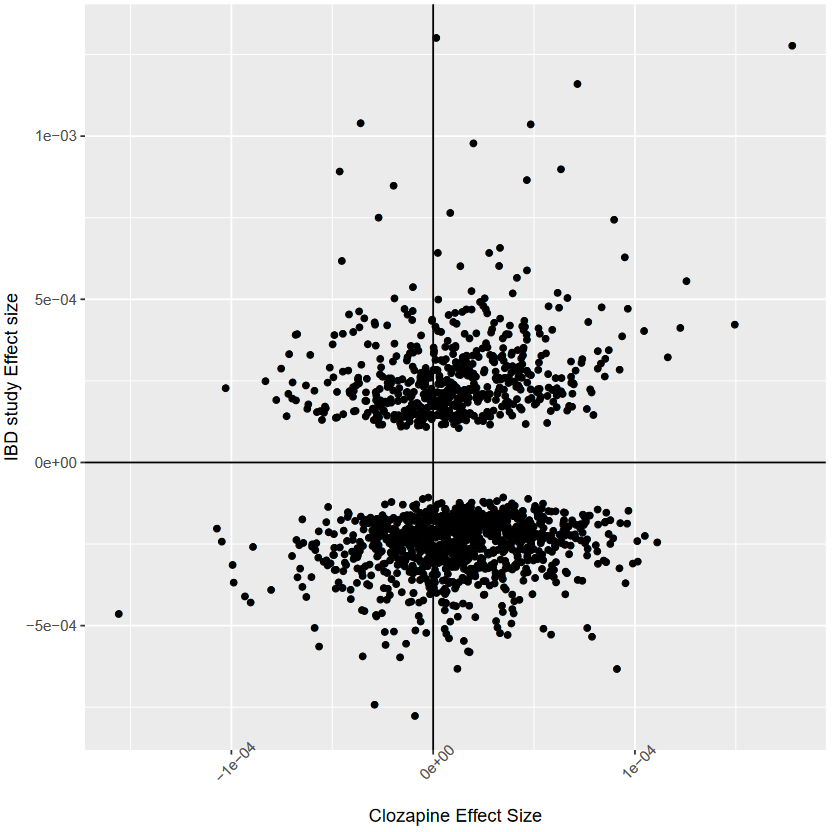

**Supplementary Figure 5. Overlap between the 1591 DMPs assayed in Lin et al., (2023) analysis of anti-TNF treatment in IBD and our analysis of DMPs associated with time on clozapine**.

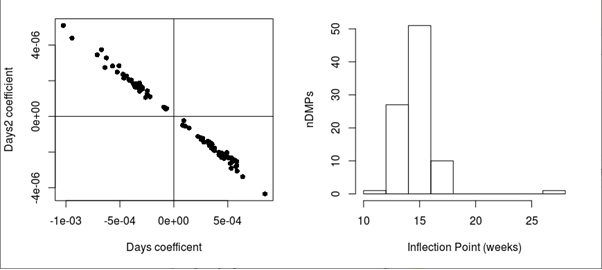

**Supplementary Figure 6. Inflection points for** **discovery DMPs associated with time on clozapine in the non-linear model.** On the left is a scatterplot comparing the estimated coefficients for the days (x-axis) and days^2^ (y-axis) on clozapine terms from the quadratic model for all discovery DMPs. On the right is a histogram showing the distribution of the inflection points for discovery DMPs associated with time on clozapine in the non-linear model.
